## Supplemental 1 for "Cohort Study Protocol of the Brazilian Collaborative Research Network on COVID-19: strengthening WHO global data"

### Description of the analysis objectives

#### Objective 1. Description of socio-demographic and clinical characteristics

Socio-demographic and clinical characteristics will be described for the population and pre-specified subgroups. Table 3 presents the description of the study variables with their respective descriptive statistics. Tables and graphs will be used to present the results.

**Table 3.** Study variables and their values or categories and numerical summary

| **Variable** | **Numerical summary** |
|---|---|
| Age (continuous and categorical)<br>< 18 years old<br>0-4 years<br>5-13 years<br>14-17 years<br>18-45 years<br>46-65 years<br>66-75 years<br>> 75 years old | mean (SD) or median (IQR), when continuous and n/N (%), when categorical |
| Gender<br>Male, female<br>If female, pregnancy status | n/N (%) |
| Anthropometry and vital signs on | |
| admission Temperature (continuous and categorical > 37.5ºC) | mean (SD) or median (IQR), when |
| Heart rate (continuous and categorical > 100 p/ min in adults and adolescents; > 120 in children up to 12 years) | continuous and n/N (%), when categorical mean (SD) or median (IQR), when continuous and n/N (%), |
| Respiration rate (continuous and categorical > 20 r/min in adults and adolescents; > 40 in children up to 12 years) | when categorical<br><br>mean (SD) or median (IQR), when continuous and n/N (%), when |
| Blood pressure (continuous and categorical systolic > 140 mmHg in | categorical |
| adults) Oxygen saturation (continuous and categorical: <90%, <94%) | mean (SD) or median (IQR), when continuous and n/N (%), when categorical mean (SD) or median (IQR), when continuous and n/N (%), when categorical |

| | |
|---|---|
| Oxygen saturation in room air/oxygen therapy (categorical) | |
| Glasgow Coma Scale | mean (SD) or median (IQR) |
| Height (cm) | mean (SD) or median (IQR) |
| Weight (kg) | mean (SD) or median (IQR) |
| BMI (continuous: kg/m$^2$ and categorical: ≥ 30;<br>≥ 40) | mean (SD) or median (IQR), when continuous and n/N (%), when categorical |
| Obesity (BMI ≥ 30) | n/N (%) |
| Chronic conditions<br>Chronic heart disease (not hypertension); Diabetes; Hypertension; Current smoker; Chronic lung disease; Tuberculosis; Asthma; Asplenia; Chronic kidney disease; Malignant neoplasm; Chronic liver disease (categorical Yes/No); Other (open text field) | n/N (%) |
| Pre-admission or chronic medications<br>ACE inhibitors; ARBs; NSAIDs; Antiviral: (if yes, which: chloroquine/hydroxychloroquine, azithromycin, kaletra [lopinavir-ritonavir], favipiravir, Other) | n/N (%) |
| Signs and symptoms on admission and during hospitalization | n/N (%) |
| Laboratory examinations on admission and<br>during hospitalization<br>Hemoglobin (g/L); Creatinine (mmol/L); White blood cell count (x10g/L); Sodium (mEq/L); Hematocrit (%); Potassium (mEq/L); Platelets (x10g/L); Procalcitonin (ng/mL); APTT/PT; CRP (mg/L); PT (seconds); LDH (U/L); INR; Creatine kinase (U/L); ALT/SGPT (U/L); | mean (SD) or median (IQR) |

| | |
|---|---|
| Troponin (ng/mL)(mEq/L); Total bilirubin (µmol/L); ESR (mm/hr); AST/SGOT (U/L); D-dimer (mg/L); Urea (mmol/L); Ferritin (ng/mL); Lactate (mmol/L) | |
| Treatments on admission and during hospitalization<br>Antiviral (If Yes, which: Ribavirin, Lopinavir/Ritonavir, Neuraminidase Inhibitor, Interferon alpha, Interferon beta, Other; If Other, specify); Corticosteroids (If Yes, route: Oral, Intravenous, Inhaled; If Yes, specify agent; If Yes, provide maximum daily dose);<br>Antibiotic; Antifungal agent; Antimalarial agent (If Yes, specify); Experimental agent (If Yes, specify); NSAID; ACE inhibitors; ARBs; Systemic anticoagulation | n/N (%) |
| Examination of the pathogen performed during this disease episode<br>Influenza virus (If Positive, specify type); Coronavirus (If Positive, specify type; If Other, specify); Other respiratory pathogen (If Positive, specify type); HIV<br>Hospitalization in ICU or High | n/N (%) |
| Dependency Unit | n/N (%) |
| Respiratory and intensive care interventions<br>Oxygen therapy; Invasive ventilation (any); Extracorporeal support (ECMO); Liver dysfunction; Pronation; Renal replacement therapy (RRT) or dialysis; Inotropics/vasopressors | n/N (%)<br>n/N (%) |

| | |
|---|---|
| Hospital outcome<br>Discharge alive, Hospitalized, Transferred to another unit, Death, Palliative discharge, Unknown | n/N (%) |
| Complications during hospitalization<br>Shock; Bacteremia; Convulsion; Bleeding; Meningitis/encephalitis; Endocarditis; Anemia; Myocarditis/pericarditis; Cardiac arrhythmia; Acute kidney injury; Cardiac arrest; Pancreatitis; Pneumonia; Liver dysfunction; Bronchiolitis; Cardiomyopathy (If Yes, specify) | n/N (%) |
| Time-to-event outcomes Symptom onset (day of disease on admission): days between first symptom and admission Length of stay<br>Length of stay in ICU<br>Days from hospital admission to ICU admission<br>Days from hospital admission to transfer or death (reported as total, for survivors and non-survivors) | median (IQR), mean (SD), survival curve (Kaplan-Meier) |

**Objective 2. Characterization of access profiles and hospital resource utilization patterns**

The characteristics signs/symptoms, laboratory tests on admission, presence of chronic diseases, treatments received, and outcomes will be described for the population and pre-specified subgroups.

**Objective 3. Association between clinical/socio-demographic characteristics and outcomes**

Measures of association, such as *odds ratio* (OR), risk ratio (RR) or *hazard ratio* (HR), crude and adjusted for potential confounding variables will be estimated and 95% confidence intervals (95% CI) will be reported. Logistic regression (or Poisson models with robust variance) will be fitted for dichotomous outcomes. Cox proportional hazards models will be fitted for time-to-event outcomes. The assumptions of the regression models will be evaluated using goodness of fit statistics (likelihood and variance analysis) and residual analysis. The set of adjustment variables will be specified primarily by building theoretical models using causal diagrams (*directed acyclic graphs* - DAGs) [5,6].

**Objective 4. Sources of inequities**

The distribution of socio-demographic, clinical, and outcome characteristics will be described and compared between pre-specified subgroups. Crude association measures (OR, RR or HR, depending on the nature of the outcome) adjusted for potential confounding variables will be estimated and

95% CIs will be reported. The set of adjustment variables will be primarily specified by building theoretical models (DAGs).
